# Factors Associated with Non-and Incomplete Vaccination Among Children Aged 12-23 Months in Gindhir Rural District, Southeast Ethiopia: A Multinomial Analysis

**DOI:** 10.1101/2024.04.19.24306059

**Authors:** Geremew Tsegaye Mulugeta, Desta Hiko Gemeda, Abraham Lomboro Dimore, Tihitian Yohannes Getu, Adamu Zerihun Gelaw, Adane Woldeab Doyamo

**Author notes:** Corresponding author: Geremew Tsegaye.

## Abstract

**Background:** Childhood immunization is one of the most successful public health interventions that prevent children’s morbidity and mortality from Vaccine Preventable Diseases (VPDs). Despite this, achieving high vaccination coverage is still one of the pressing public health problems globally, particularly in low-and middle-income countries.

**Objective:** This study aims to identify factors associated with non- and incomplete vaccination status among children aged 12-23 months in Gindhir rural district, East Bale zone, Southeast Ethiopia.

**Methods:** A cross-sectional study supplemented with a qualitative study was conducted in the Gindhir district from April 25-June10/ 2021. A total of 563 mothers of children 12-23 months participated. Study participants were selected using computer-generated random numbers. The sample was proportionally allocated to the size of children 12-23 months in each kebele. An administered structured questionnaire was used to collect data using face-to-face interviews. Epi– data version 3.1 was used for data entry and SPSS version 25 software was used for data analysis. Bivariate and Multinomial logistic regression analysis was used to identify the determinants of vaccination status at a P value of less than 0.05. Seven in-depth interviews and two focused group discussions were conducted and analyzed manually by coding.

**Results:** -Totally 563 mothers/caregivers with children aged 12-23 months participated with a 96.6% response rate. Of the 563 children included in the study, 307 (54.5% [95%CI: 50.1-58.8]) were fully vaccinated, 142(25.5% [95%CI:21.7-29.1]) were incompletely vaccinated and 114 (20.2% [95%CI: 16.7-23.8]) were not vaccinated at all. Home delivery [3.46 (95%CI: 1.27-9.40)], absence of nearby health facility[4.84(95% CI: 1.75-13.39)], lack of transportation incur cost [2.75(95%CI:1.06-7.14)], mothers/caregivers negative perceived benefits of child vaccination [2.69(95%CI: 1.35-5.37)], mothers/caregivers poor knowledge about VPDs [2.78(95%CI: 1.14-5.30)] and vaccination schedule [5.54(95%CI: 2.74-11.22)], and negative attitude towards vaccination[2.01(95%CI: 1.01-3.98)] were identified as independent predictors of non-vaccination. While home delivery [6.85 (95%CI: 1.69-27.79)], lack of provision of counselling by health workers [2.19(95%CI: 1.13-4.27)], mothers/caregivers’ poor knowledge about VPDs [2.71(95%CI: 1.60-4.58)] and vaccination schedule [3.30(95%CI: 1.90-5.74)], and attitude towards vaccination [2.53(95%CI: 1.47-3.38)] were significantly associated with incomplete vaccination status.

**Conclusion:** Designing and implementing public health interventions tailored to locally identified problems is vital to narrow the observed variation in childhood vaccination status.

## Introduction

Childhood immunization is one of the most successful public health interventions that prevent children’s morbidity and mortality from Vaccine Preventable Diseases (VPDs). Globally, immunization saves 2-3 million of life per year. However, VPDs still account worldwide for over 2 million under-five deaths annually, the majority of them being from countries in sub-Saharan Africa^(1)^. Synchronously, achieving high vaccination coverage is still one of the pressing public health problems globally, particularly in low and middle-income countries. According to World Health Organization (WHO), global vaccination coverage dropped from 86% in 2019 to 81% in 2021. Furthermore, it was reported that in 2021, an estimated 25 million children under the age of 1 year did not receive basic vaccines, and the number of completely unvaccinated children increased by 5 million since 2019^(2)^.

In African region, almost 8.5 million children were unvaccinated or incompletely vaccinated, which is higher compared to other regions. For example, DTP3 coverage was below 60% in nine African countries in 2018. In addition, there were 19.4 unvaccinated children in this region, of which about 8.6 million (44%), of them live in 16 countries where polio is endemic and affected by conflict like Nigeria and Ethiopia ^(3)^. Additionally, 9.4 million of the 19.7 million children who are not fully protected against VPDs (without DPT3) resides in Africa, particularly in Nigeria, the Demographic Republic of the Congo, Ethiopia and Angola^(4)^.

Even though different vaccination strategies were implemented to expand lifesaving vaccine, VPDs are still account worldwide for over 2 million under-five deaths annually, the majority of them being from countries in sub-Saharan Africa^(1)^. Despite this fact, achieving high vaccination coverage is still one of the pressing public health problems globally, particularly in low and middle-income countries^(2)^. In Africa about more than half a million children die from VPDs every year, accounting for 58% of global deaths. Furthermore, many African countries affected by outbreaks of VPDs such as pneumococcal disease, yellow fever, measles, and rotavirus ^(5)^.

Ethiopia launched the Expanded Program on Immunization (EPI) in 1980 to reduce maternal and child morbidity and mortality. Since then, EPI has been one of the core priorities in the country’s past Health Sector Development Programs (HSDPs) and current Health Sector Transformation Plan (HSTP). Vaccinations are routinely given at the country’s static, outreach, and mobile health facilities. Reaching Every District (RED) and Sustainable Outreach Services (SOS) approaches were also introduced in 2003. Additionally, to address the immunization inequity and increase coverage, the Periodic Intensification of Routine Immunization (PIRI) has been implemented since 2018 in selected poor-performing woredas of agrarian and pastoral regions. Moreover, other strategies to increase immunization coverage such as Child Health Day events, intensified outreaches, and pulse campaigns have been implemented. Furthermore, the country has mobilized women’s development armies or volunteers, health extension workers, and health facilities to deliver immunization services^(6)^.

However, achieving a national vaccine plan is still one of the public health challenges. According to a study that analyzed data from the Ethiopian Demographic and Health Survey (EDHS) the overall prevalence of fully vaccinated, partially vaccinated and non-vaccinated children aged 12-23 months was 35%, 49% and 16%, respectively^(7)^. Similarly, Ethiopian Mini Demographic and Health Survey (EMDHS) 2019 reported that only 4 in 10 children (43%) were fully vaccinated, and close to 2 in 10 children (19%) were non-vaccinated. Likewise, there is regional variation in coverage of all basic vaccinations, which is highest in Addis Ababa (83%) and lowest in Afar (20%) ^(8)^.

To achieve the full benefits of vaccination, understanding barriers to childhood vaccination is essential. The interventions should also be tailored to the locally identified barriers. Studies conducted previously on childhood vaccination have reported that maternal education, socioeconomic status, antenatal care, place of delivery, sex of a child, postnatal care, media exposure, perceptions of vaccination, place of residence and cold chain management were contributing factors to childhood vaccination status ^(7,9–11)^.

Even though there are studies conducted on childhood vaccination status in Ethiopia, the studies have certain limitations. First, almost all the previous studies were conducted on full or incomplete vaccination status, which did not address the issue of non-vaccination. But as far as our knowledge, literature categorizes the vaccination status of children as fully vaccinated, partially vaccinated and non-vaccinated. While interventions needed to solve non-and incomplete vaccination status are not the same.

Hence, studies that identify predictors of childhood vaccination using the three-way categories of full vaccination, incomplete vaccination and non-vaccination are crucial. Similarly, previous studies conducted in Ethiopia on childhood vaccination status have focused on either individual vaccine coverage or full and incomplete vaccination, and none of these studies were conducted to identify the predictors of full, incomplete and non-vaccination status. Even though, there was a study conducted on child hood vaccination status and associated factors in the EHDS, categorizing in to fully vaccinated, incomplete and non-vaccination, the study used secondary data from EDHS 2016 that was the population-based sample data. And, some important indicators such as individual (child) level, maternal health services utilization related predictors, their knowledge about vaccine and vaccination schedule, attitude towards vaccination, and healthcare system-related factors were not assessed. Therefore, the current study tried to identify the predictors of non- and incomplete vaccination status among children aged 12-23 months in Gindhir district, East Bale zone, Southeast Ethiopia.

## Methods and Materials

### Study Setting and Period

The study was conducted in Gindhir rural district Bale Zone, Southeast Ethiopia from April 25, 2021to June 10, 2021. Gindhir rural districts one of the districts of Bale zone and is geographically located at 7ₒ8’0’’North, 40’42’’0’’East. It is bordered on the south by the Gastro River that separates it from Goro, on the west by Sinana on the northwest by Gasera and Gololcha, on the Northeast by Sewena, and the East by Raytu. Gindhir rural districts located at 502.1km distance from Addis Ababa. The total population of the district is 148,886 according to demographic and health survey in 2020 and about 4466 were children 12-23-month age. The district has an estimated population density of 59.8 people per square kilometers. The district has 32 kebele (lowest administrative unit in Ethiopia), with eight health centers and 32 health posts for providing primary health care services including vaccination.

### Study design

Community-based cross-sectional study triangulated with a qualitative data collection method was employed from April 25-June10/ 2021. The qualitative study involved a purposively selected seven key informant interviews and two Focused Group Discussions (FGD).

#### Population

The source population for this study was all mothers/caregivers with children aged 12-23 months in the Gindhir rural district, while the study population was mothers/caregivers with children aged 12-23 months who resided in the district for at least six months in selected kebeles and available during the study period. Mothers/caregivers aged 15 and above years were included and mothers/caregivers who can’t provide information for different reasons such as serious illness were excluded from the study.

#### Sample Size Determination and Sampling Procedure

Sample size was determined using a single population proportion estimation formula by considering the following assumptions: prevalence of non-vaccination status (19%) from EDHS 2019 (8), 95% Confidence Interval (CI), margin of error (4%), design effect (1.5), and non-response rate (5%). And the determined final sample size was 583.

Regarding sampling technique, we used a multi-stage sampling technique. Gindhir rural district has 32 rural kebeles and we selected 10 kebeles randomly. Before the actual data collection survey was conducted in the selected kebeles to get the list of households who have children aged 12-23 months. The survey was conducted by twenty leaders of the health development army who were able to read and write the local language. Then sampling frame having the list of children aged 12-23 months for each kebeles was prepared separately. The total sample size was proportionally allocated to each kebeles depending on the total number of children aged 12-23 months in each kebeles. Finally, individual mothers/caregiver with children aged 12-23 months was recruited using a computer-generated simple random sampling technique as show Figure 1 below.

**Figure 1:**
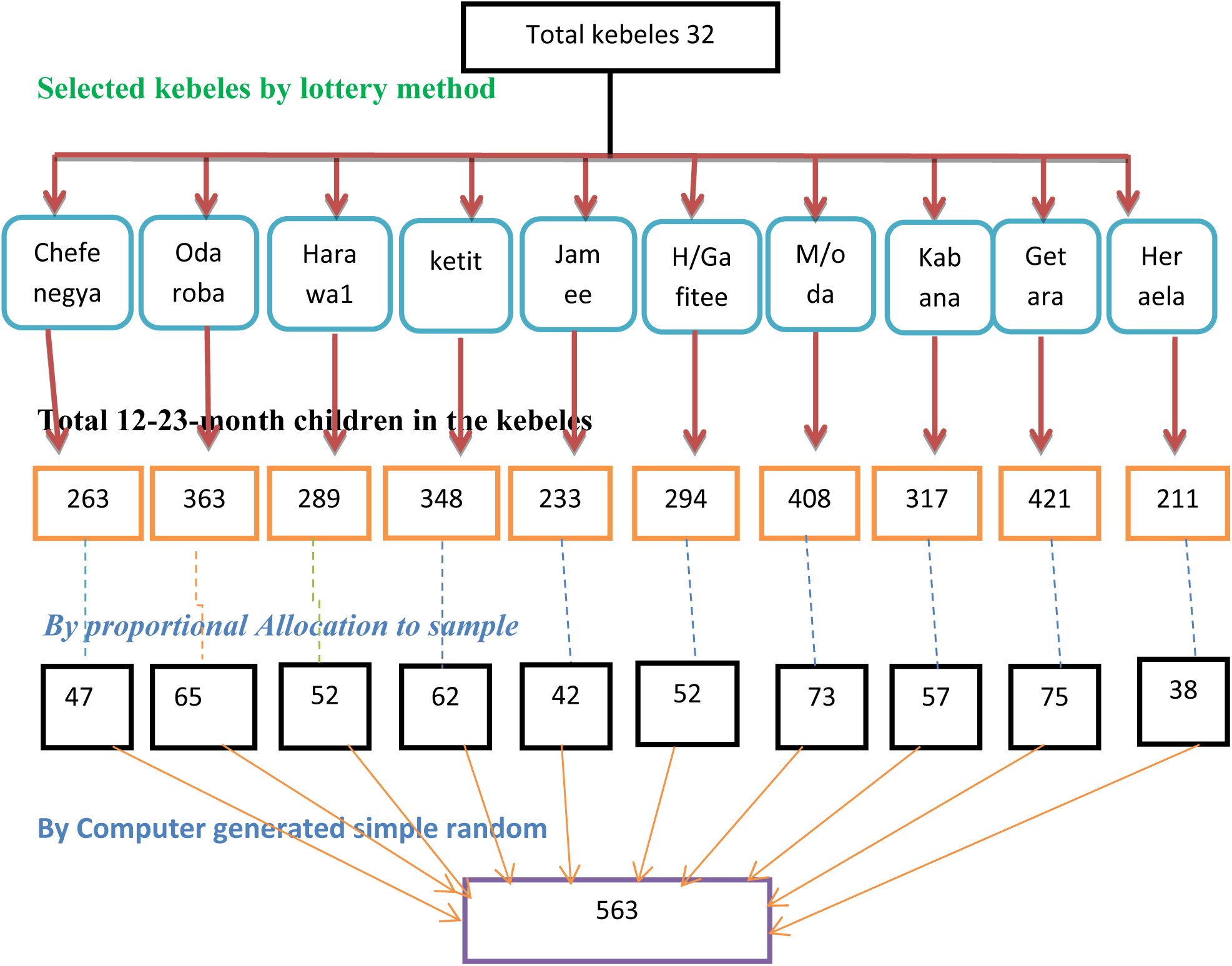
Schematic presentation, showing multi stage sampling of the study

For the qualitative part, a purposive sampling technique was employed to select the key informants and participants in FGD. The key informants were the district EPI focal person, three EPI focal persons from health centers with high non –and/or incomplete vaccination coverage within their catchment kebele, and three health extension workers from health posts. Additionally, two FDGs (each consisting of eight participants) were conducted with mothers of non- and incompletely vaccinated children of the 12–23-month age group. Both key informants’ interviews and FGD were continued until information saturation was reached, meaning the investigator agreed that there was redundancy in the responses and no new idea emerging.

#### Data Collection Procedure

We prepared an interviewer-administered structured questionnaire for a quantitative study after reviewing previously conducted studies ^(7,9–11)^. The questionnaire consisted of six sections including; socio-demographic, child immunization history, health service-related, knowledge about VPDs and vaccination schedule, attitude towards vaccination, reason for non-vaccination and maternal health services utilization-related questions. Source of information for vaccination was immunization card, history from mothers/caregivers, and completion certificates. Data were collected through face-to-face interviews with the mothers/caregivers and a review of the immunization cards. Data were collected by ten experienced nurses under the supervision of two Health officers. If there were two eligible children in one household, information was taken only for the youngest one because it provides the most recent information.

For the qualitative study, the key informants were interviewed using a semi-structured interview guide with a flexible probing technique and FGD guidelines were used to conduct FGD with mothers of 12-23 months children. The key informant interview was performed by the experienced interviewer, while FGD was done with a team consisting of one modulator, one note taker and one recorder including the principal investigator. Before conducting the interview, an explanation and elaboration of the need to do the key interview and FGD was made and the participants were asked for their willingness to participate. The Interviews were conducted at the workplaces of the EPI focal persons and health extension workers and the FGD was conducted at the kebele level. An interview takes an average of 30 minutes and FGD 1 hour and 30 minutes. All the information was tape - recorded and field notes were taken.

#### Measurements and Operational Definition

Knowledge of mothers/caregivers about VPDs was measured by asking the participants to mention the list of vaccine-preventable diseases they know. A score of 1 was given for the correct answer and 0 for the incorrect answer. Those mothers/caregivers of children aged 12-23 months who mentioned the list of less than nine (75%) VPDs correctly were categorized as having poor knowledge about VPDs. Similarly, knowledge about vaccination schedules was measured by asking the participants to state the eight vaccination schedules. The correct answer was score 1 and the incorrect answer scored 0. Then, participants who mentioned at least five schedules correctly were considered to have good knowledge about vaccination schedules, otherwise poor.

Mothers/caregivers’ attitude towards vaccination is assessed using twelve questions on a five-point Likert-type scale, where 1= strongly disagrees and 5= strongly agree. The questions included both positively and negatively worded statements. Five items have been negatively worded, which requires reverse coding. Internal consistency was checked using reliability statistics with Cronbach’s α=0.85 during the pretest. The total score was computed and participants who scored less than the mean score were considered as having a negative attitude towards vaccination.

### Non-vaccinated

Non-vaccinated is defined as a child aged 12-23 months who did not receive any vaccine recommended by the national EPI schedule before his/her birthday.

### Incomplete vaccinated

A child aged 12-23 months who received fewer than all doses recommended in the national immunization schedule, but at least one immunization.

### Fully vaccinated

A child aged 12-23 months who received all currently recommended vaccines (one dose of BCG, three doses of Penta-valent, three doses of OPV, excluding which provided at birth, three doses of PCV, two doses of Rotavirus vaccine, and one dose of measles vaccine) any time before the data collection was taken as fully vaccinated ^(12)^.

### Perceived benefits of vaccine

Mothers/ caretakers who were aware that the vaccine could prevent the disease for which the child is vaccinated were considered as having perceived benefits of the vaccine ^(38)^.

### Data Quality Management

We provided two-day training for data collectors and supervisors on the objectives of the study, sampling procedure and approach during data collection. Translation of the data collection tool from the English language to the Afaan Oromo language and back-translated to the English language to ensure consistency was performed by language experts. Before data collection, a pre-test of the data collection tool was done on 5% of the sample size in the Sinana district. When the participants were not available on the first visit, an additional three visits to their homes were made to minimize the non-response rate. The data collection process was supervised and checked for completeness daily by supervisors. The principal investigator also closely followed and checked data for completeness on a daily basis.

### Statistical Analysis

Data were entered and cleaned on Epi-data version 3.1 software and exported to SPSS version 26 software for analysis. Descriptive analysis such as mean and standard deviation for continuous variables, and frequency and percentage for categorical variables were used to summarize and explain characteristics of study variables. The dependent variable was nominated into non-vaccinated, incompletely vaccinated and fully vaccinated. Fully vaccinated was used as a reference variable for comparison. Bivariate analysis was employed to see the association between each independent variable and dependent variable. Variables with a p-value of less than 0.25 were considered as candidates for the final model. Multinomial logistic regression was used to determine the independent predictors of non-vaccination and incomplete vaccination status. The strength of association was assessed using Adjusted Odds Ratio (AOR) with their respective 95% confidence intervals. The model was tested and adequately fitted at Pearson Chi-square of 438.8 and p-value of 0.01. The model goodness of fit is also significant at a p-value of 0.182 and Pseudo R-squared of 0.788. The classification capacity of the model is 75 per cent.

### Ethical Consideration

Ethical approval was obtained from Jimma University’s ethical review committee with **Ref. No. IRB-00018/2020,** and submitted to East Bale Zone Health Department. An official support letter was written from the Zonal Health Department and given to the Gindhir health office. After explaining the study’s purpose, procedure, potential benefits and risks, and their right to participate or not, written informed consent was obtained from mother / care giver of the study participants. Data were collected anonymously; information obtained was used for study purposes only and not disclosed to ensure confidentiality. Those mothers/caretakers of non-vaccinated or incompletely vaccinated children were advised to vaccinate their children.

## Results

### Socio Demographic Characteristics of Participants

A total of 563 mothers/caretakers with their children aged 12-23 months have participated in this study which makes a response rate of 96.6%. Nearly one-third (30.2%) of mothers were found within the age category of 35-49 years. The mean (SD) age of the study participants is 29.99 (±8.32). Regarding the occupational and religious status of the study participants, 342(60.6%) and 357(63.4%) of them were housewives and Muslim religious followers, respectively. From the total, more than three-fourths (76.7%) of the study participants were residing in rural areas of the district. Regarding the educational status of the mothers about 219 (39%) of the mothers have no formal education. Of the total participants, one-third (33%) had a large family size of 5-10 members and about 173(30.7%) of them experienced seasonal migration within 2 years for farming and grassing **(Table 1**).

**Table 1:** Socio demographic characteristics of study participants for the study on factors associated with non-and partial vaccination among children aged 12-23 months in Gindhir District, Southeast Ethiopia 2021.

| Variable | Category | Frequency | Percent |
| --- | --- | --- | --- |
| Child caretaker | Mother | 291 | 51.7 |
|  | Child guard | 72 | 12.8 |
|  | Grandmother/father | 200 | 35.5 |
| Marital Status | Single | 77 | 13.7 |
|  | Married | 421 | 74.8 |
|  | Widowed | 48 | 8.5 |
|  | Divorced | 17 | 3.0 |
| Educational status | No formal education | 219 | 38.9 |
|  | Primary education | 254 | 45.1 |
|  | Secondary and above | 90 | 16.0 |
| Religion | Muslim | 357 | 63.4 |
|  | Orthodox | 161 | 28.6 |
|  | Protestant | 45 | 8.0 |
| Family size | 1 – 5 | 376 | 66.8 |
| | $\geq 6$ | 187 | 33.2 |
| Occupational status | House wife | 342 | 60.6 |
|  | Farmer | 56 | 9.9 |
|  | Merchant | 46 | 8.2 |
|  | Government employee | 53 | 9.4 |
|  | Others | 67 | 11.9 |
| Seasonal migration | Yes | 173 | 30.7 |
| within last two years | No | 390 | 69.3 |
| Residence | Rural | 432 | 76.7 |
|  | Urban | 131 | 23.3 |
| Age of the mother/caretaker | 15-19 | 68 | 12.1 |
|  | 20-24 | 101 | 17.9 |
|  | 25-29 | 117 | 20.8 |
|  | 30-34 | 107 | 19.0 |
|  | 35-49 | 170 | 30.2 |

### Vaccination Status of the Children

In this study data on child vaccination was collected based on child immunization cards 372(66%), medical records 77(14%) and mothers’ recall 114(20.2%). Out of 563 mothers/caretakers with children aged 12-23 months, 307(54.5% [95%CI: 50.1-58.8]) of them were fully vaccinated, 142(25.5% [95%CI: 21.7-29.1]) incomplete vaccinated and 114(20.2% [95%CI: 16.7-23.8]) of them were non-vaccinated by routine immunization program.

There was a high difference between children’s vaccination status regarding their socio-demographic characteristics. The majority of non-vaccinated 90(79%) and incompletely vaccinated 115(81%) children were residing in rural areas. Similarly, a high proportion of children with non-vaccinated and incompletely vaccinated were observed among mothers/caregivers with primary educational status, 50(44%) and 63(44%), respectively. Regarding occupational status, the majority of children with mothers/caregivers of housewives were non-vaccinated and incompletely vaccinated 64(56.1%) and 84(59.2%), respectively. Non-vaccinated children were highly noticed among children with male sex 58(50.9%) whereas incompletely vaccinated children are equal for both sexes (50%). Moreover, high proportion of non-vaccinated 78(68.4%) and incomplete vaccinated 86(60.6) children were within birth order of 1-3 **(Table 2)**.

**Table 2:** Characteristics of sample by vaccination status in Gindhir rural districof East Bale Zone, 2021.

| Variable | Category | Non-Vaccinated N(%) | In completely Vaccinated N (%) | Fully vaccinated N (%) |
| --- | --- | --- | --- | --- |
| Vaccination status |  | 114(20.2) | 142(25.5) | 307(54.5) |
| Place of residence | Urban | 24(21) | 27(19) | 80(26) |
|  | Rural | 90(79) | 115(81) | 227(74) |
| Marital status of mother/caregiver | Single | 26(22.8) | 24(16.7) | 27(8.8) |
|  | Married | 77(67.5) | 95(66.9) | 249(81.1) |
|  | Widowed | 10(8.8) | 19(13.4) | 19(6.2) |
|  | Divorced | 1(0.9) | 4(2.8) | 12(3.9) |
| Educational status of mother/caregiver | No formal education | 45(40) | 53(37) | 121(39) |
|  | Primary education | 50(44) | 63(44) | 141(46) |
|  | Secondary and above | 19(17) | 26(18) | 45(15) |
| Occupation status of the mother | House wife | 64(56.1) | 84(59.2) | 193(62.9) |
|  | Farmer | 13(11.4) | 15(10.6) | 28(9.1) |
|  | Merchant | 12(10.5) | 11(7.7) | 23(7.5) |
|  | Government employee | 14(12.3) | 12(8.5) | 27(8.8) |
|  | Others | 11(9.6) | 20(14.1) | 36(11.7) |
| Family size | 1-5 | 70(61.4) | 89(62.9) | 190(61.9) |
| | $\geq 6$ | 44(38.6) | 53(37.3) | 117(38.1) |
| Sex of child | Male | 58(50.9) | 71(50.0) | 158(51.5) |
|  | Female | 56(49.1) | 71(50.0) | 149(48.5) |
| Birth order | 1-3 | 78(68.4) | 86(60.6) | 203(66.1) |
| | $\geq 4$ | 36(31.6) | 56(39.4) | 104(33.9) |
| Seasonal Migration | No | 67(58.8) | 99(69.7) | 224(73.0) |
|  | Yes | 47(41.2) | 43(30.3) | 83(27.0) |

### Knowledge About Vaccine and Attitude Towards Vaccination

In this study mothers/caregiver’s knowledge about vaccines and vaccination schedules was measured by asking them to mention the list of 12 VPDs and eight vaccination schedules they know, respectively. Accordingly, 75(65.8) of non-vaccinated and 83(58.5) of incompletely vaccinated children’s mothers/caregivers had poor knowledge about VPDs. Correspondingly, nearly four-fifths (78.9%) of non-vaccinated and more than three-fifths (63.4%) of incompletely vaccinated children’s mothers/caregivers had poor knowledge about vaccination schedules. On the other hand, about three-fifths (61.4) of non-vaccinated and more than three-fifths (62.7) of incompletely vaccinated children’s mothers/caregivers had negative attitudes towards vaccination **(Table 3).**

**Table 3:** Knowledge about vaccine and attitude towards vaccination of study participants in Gindhir district, East bale zone, Ethiopia, 2021.

| Variable | Category | Non-vaccinated<br>N (%) | Incompletely vaccinated N (%) | Fully vaccinated N (%) |
| --- | --- | --- | --- | --- |
| Knowledge about vaccine preventable diseases | Poor | 75(65.8) | 83(58.5) | 191(62.2) |
|  | Good | 39(34.2) | 59(41.5) | 116(37.8) |
| Knowledge about vaccination schedule | Poor | 90(78.9) | 90(63.4) | 108(35.2) |
|  | Good | 24(21.1) | 52(36.6) | 199(64.8) |
| Attitude towards vaccine | Negative | 70(61.4) | 89(62.7) | 93(30.3) |
|  | Positive | 44(38.6) | 53(37.3) | 214(69.7) |
| Perceived benefits of vaccine | No | 74(64.9) | 54(38.0) | 77(25.1) |
|  | Yes | 40(35.1) | 88(62.0) | 230(74.9) |

**Table 4:** Health facility and service-related characteristics of study participants in Gindhir rural district east bale zone, 2021.

| Variable | Category | Non-Vaccinated<br>N (%) | Incompletely Vaccinated<br>N (%) | Fully vaccinated<br>N (%) |
| --- | --- | --- | --- | --- |
| Presence of health facility for EPI service | No | 48(42.1) | 20(14.1) | 20(6.5) |
|  | Yes | 66(57.9) | 122(85.9) | 287(93.5) |
| Type of HF | Health post | 45(39.5) | 102(71.8) | 161(52.4) |
|  | Health center | 38(33.3) | 40(28.2) | 146(47.6) |
| Distance from the nearest health facility | <20 minutes | 42(36.8) | 73(51.4) | 173(56.4) |
|  | 21-40minutes | 27(23.7) | 42(29.6) | 87(28.3) |
|  | ≥41 minutes | 45(39.5) | 27(19.0) | 47(15.3) |
| Means of transportation to health facility | By car/motor | 65(57.0) | 78(54.9) | 57(18.6) |
|  | Horse | 24(21.1) | 18(12.7) | 36(11.7) |
|  | Walk | 25(21.9) | 46(32.4) | 214(69.7) |
| Transportation incur cost | No | 91(79.8) | 90(63.4) | 97(31.6) |
|  | Yes | 23(20.2) | 52(36.6) | 210(68.4) |
| Health workers provide advice on vaccination | No | 42(36.8) | 43(30.3) | 38(12.4) |
|  | Yes | 72(63.2) | 99(69.7) | 269(87.6) |
| Health facility provide outreach vaccine service | No | 98(86.0) | 112(78.9) | 242(78.8) |
|  | Yes | 16(14.0) | 30(21.1) | 65(21.2) |
| Health facility trace missed child | No | 7(6.1) | 27(19.0) | 118(38.4) |
|  | Yes | 107(93.9) | 115(81.0) | 189(61.6) |

### Health Facility Related Characteristics

Regarding access to health facilities that provide immunization services, more than two-fifths (42.1) of non-vaccinated and 20(14.1) of incompletely vaccinated children live in areas where no nearby health post or health center is present. On the other hand, non-vaccination and incomplete vaccination were high among children who get vaccination services from health post 45(39.5) and 102(71.8), respectively. Nearly two-fifths (39.5) of non-vaccinated and 27 (23%) of incompletely vaccinated children reside in areas greater than 41 minutes from the nearest health facility that provides vaccination service. The highest proportion of mothers/caregivers with non-vaccination 98(86.0) and incompletely vaccinated 112(78.9) children aged 12-23 months reported that health facilities in their area did not provide outreach vaccination services (Table4).

### Maternal/caregivers Health Service Utilization

Regarding maternal/caregiver health service utilization high proportion of non-vaccinated children were observed among those who gave birth at home 46(40.4%), not started using family planning at six weeks 60(57%), no PNC visits 91(79.8%), and not trained on child care94(82.5%) and vaccination96(84.2%). Similarly, the majority of incompletely vaccinated children were seen among mothers/caregivers with the place of delivery at health post 55(38.7%), not trained on child care 94(66.2%) and vaccination 98(69%) **(Table 5).**

**Table 5:** Maternal/caregivers health service utilization related characteristics of study participants in Gindhir rural district east bale zone, 2021.

| Variable | Category | Non-Vaccinated<br>N (%) | Incompletely<br>Vaccinated<br>N (%) | Fully<br>vaccinated<br>N (%) |
| --- | --- | --- | --- | --- |
| ANC follow-up during pregnancy | No | 44(38.6) | 48(33.8) | 43(14.0) |
|  | Yes | 70(61.4) | 94(66.2) | 264(86.0) |
| Frequency of ANC follow-up | 1-3 visits | 45(39.5) | 56(39.4) | 137(44.6) |
|  | ≥ 4 | 69(60.5) | 86(60.6) | 170(55.4) |
| Place of delivery | Home | 46(40.4) | 46(32.4) | 27(8.8) |
|  | Health post | 30(26.3) | 55(38.7) | 166(54.1) |
|  | Health center | 34(29.8) | 29(20.4) | 56(18.2) |
|  | Hospital | 4(3.5) | 12(8.5) | 58(18.9) |
| PNC visit | No | 74(64.9) | 67(47.2) | 96(31.3) |
|  | Yes | 40(35.1) | 75(52.8) | 211(68.7) |
| Mother/caregiver started using family planning at 6 weeks | No | 65(57.0) | 69(48.6) | 90(29.3) |
|  | Yes | 49(43.0) | 73(51.4) | 217(70.7) |
| Training on child health care | No | 94(82.5) | 94(66.2) | 111(36.2) |
|  | Yes | 20(17.5) | 48(33.8) | 196(63.8) |
| Training on vaccination | No | 96(84.2) | 98(69.0) | 117(38.1) |
|  | Yes | 18(15.8) | 44(31.0) | 190(61.9) |

### Factors Associated with Non-Vaccination and Incomplete Vaccination

Bivariate analysis was performed to identify candidate variables for Muti-nominal logistic regression for both non-vaccination and incomplete vaccination status using fully vaccination status as a reference category. Accordingly, age of mother/caregiver, place of residence, marital status, ANC follow-up, place of delivery, PNC visit, mothers started using family planning within six weeks, presence of nearby health facility, type of health facility present nearby, means of transportation to the health facility, paying transportation incur a cost, health workers provide counselling on vaccination, health facility trace missed child, mother/caregiver trained on child care and vaccination, perceived benefits of vaccination, mothers/caregivers knowledge about VPDs and vaccination schedule, and attitude towards vaccination were associated with both non- and incomplete vaccination at a p-value of less than 0.25. Moreover, variables such as the presence of seasonal migration of mothers/caregivers and health facilities providing outreach vaccination services were identified as candidate variables for non-vaccination at a p-value of less than 0.25. We performed a final multinomial regression analysis using these candidate variables to identify independent predictors of non-vaccination and incomplete vaccination.

In Multinomial logistic regression, home delivery[3.46 (95%CI: 1.27-9.40)], absence of nearby health facility[4.84(95% CI: 1.75-13.39)], lack of transportation incur cost [2.75(95%CI: 1.06-7.14)],mothers/caregivers negative perceived benefits of child vaccination[2.69(95%CI: 1.35-5.37)], mothers/caregivers poor knowledge about VPDs[2.78(95%CI: 1.14-5.30)] and vaccination schedule[5.54(95%CI: 2.74-11.22)], and negative attitude towards vaccination[2.01(95%CI: 1.01-3.98)] were identified as independent predictors of non-vaccination. While home delivery [6.85 (95%CI: 1.69-27.79)], lack of provision of counselling by health workers [2.19(95%CI: 1.13-4.27)], mothers/caregivers’ poor knowledge about VPDs [2.71(95%CI: 1.60-4.58)] and vaccination schedule [3.30(95%CI: 1.90-5.74)], and attitude towards vaccination [2.53(95%CI: 1.47-3.38)] were significantly associated with incomplete vaccination status **(Table 6)**.

**Table 6:** Multinomial logistic regression for factors associated with non-and incomplete vaccination among children aged 12-23 months in Gindhir district, East Bale zone, Ethiopia, 2021.

| Factors | Category | Non-versus fully vaccinated AOR (95% CI) | P-value | Incomplete versus fully vaccinated (AOR (95%CI) | P-value |
| --- | --- | --- | --- | --- | --- |
| Place of delivery | Home | 3.46 (1.27-9.40) | 0.015 | 6.85 (1.69-27.79) | 0.007 |
|  | HP | 1.30 (.59-2.88) | 0.506 | 2.98(.81-10.88) | 0.099 |
|  | HC | 1.10 (.44-2.70) | 0.833 | 3.51(.93-13.22) | 0.063 |
|  | Hospital | 1 |  | 1 |  |
| Presence of health facility for EPI service | No | 4.84(1.75-13.39) | 0.002 | 1.18(0.44-3.15) | 0.74 |
|  | Yes | 1 |  | 1 |  |
| Transportation incur cost | No | 2.75(1.06-7.14) | 0.04 | 1.57(0.74-3.34) | 0.24 |
|  | Yes | 1 |  | 1 |  |
| Health workers provide counseling on vaccination | No | 2.06(0.94-4.51) | 0.07 | 2.19(1.13-4.27) | 0.02 |
|  | Yes |  |  |  |  |
| Perceived benefits of vaccine | No | 2.69(1.35-5.37) | 0.01 | 1.07(0.61-1.88) | 0.83 |
|  | Yes | 1 |  | 1 |  |
| Knowledge about VPDs | Poor | 2.78(1.14-5.30) | <0.001 | 2.71(1.60-4.58) | <0.001 |
|  | Good | 1 |  | 1 |  |
| Knowledge about vaccination schedule | Poor | 5.54(2.74-11.22) | <0.001 | 3.30(1.90-5.74) | <0.001 |
|  | Good | 1 |  | 1 |  |
| Attitude towards vaccination | Negative | 2.01(1.01-3.98) | 0.04 | 2.53(1.47-3.38) | 0.001 |
|  | Positive | 1 |  | 1 |  |

## Discussion

This study assessed factors associated with non-and incomplete vaccination among children aged 12-23 months, in the Gindhir rural district of East Bale, Oromia region. Our results show that out of 563 children, 54.5% of them were fully vaccinated, 25.5% were incompletely vaccinated and 20.2% were non-vaccinated by routine immunization program. We used multinomial logistic regression to identify factors affecting non-and incomplete vaccination status. Factors associated with non-and incomplete vaccination were assessed separately by taking full vaccination status as a reference category. Consequently, home delivery, poor knowledge of mothers/caregivers about VPDs and vaccination schedule, and negative attitude towards vaccination were significantly associated with both non- and incomplete vaccination status. While, the absence of nearby health facilities, transportation incur costs, and negative mothers/caregivers’ perceived benefit of vaccine were associated with non-vaccination status. However, the lack of provision of counselling about vaccination was identified as an independent predictor of incomplete vaccination status only.

In this study, more than half (54.5%) of children aged 12-23 months were fully vaccinated. This is consistent with findings from studies conducted previously^(13–15)^. However, this coverage is less than the benchmark of 90% set by WHO and the national target set by the Ethiopian government in 2020^(6,16)^. Additionally, 25.5% of children were incompletely vaccinated, which means they received one or more doses of any basic vaccines, but not all of them. In the meantime, considerable proportions (20.2%) of children never received any of these basic vaccines. These findings are comparable with the findings of studies previously conducted elsewhere ^(7,9,15,17,18)^. This low coverage indicates that there is a gap in addressing all children in every district with routine childhood vaccination and challenges to achieve high vaccination coverage remain. Thus, it highlights the necessity of strengthening the implementation of RED strategies to reach and sustain high vaccination coverage to increase protection from VPDs for all children. Moreover, increasing and maintaining high vaccination coverage need to be addressed in a country and context-specific manner ^(19)^.

> Supported by qualitative finding ……… *“Many children default every month; we did not know what the reason behind why children in our catchment are defaulting from vaccination is… may be… it is due to fear of vaccine side effect or gap in information about immunization but they vaccinate their children simply by looking each other and better if we avert this trend by advising mothers but we have a gap on advising on routine childhood immunization”*.

> *Qualitative findings also support this. "We identified areas like the Abayi Zone that would benefit from outreach programs. However, we are currently unable to provide outreach services in these areas. Traveling long distances alone creates a security risk for our staff. Additionally, using motorbikes for travel is expensive, costing around 400 ETB per trip. Due to these limitations, we can no longer offer outreach services in the same way. Instead, we advise residents to visit the nearest health facility for vaccinations, but unfortunately, many do not follow through”*.

The current study found that children who were born at home were more likely to be non- and incompletely vaccinated than fully vaccinated compared to those who were born in a hospital. This result is in line with studies conducted earlier^(17,20,21)^, which reported being delivered in the family /home increases the risk of incomplete and non-vaccination. Mothers giving birth in hospitals receive crucial vaccination counselling, while those choosing home birth often have no such access, leaving parents uninformed about critical immunizations^(22)^. In addition, health workers provide the BCG and OPV0 vaccines for those children who are born at health facilities, as well as advise their mothers about subsequent vaccines due to policy matters. The finding implies that encouraging institutional delivery increases the likelihood of fully vaccinating children.

> *Supported by qualitative finding ……” Many mothers in our catchment area practice home delivery and miss focused postnatal care. During focused postnatal care, we provide information about newborn care, including childhood vaccination. Midwives advise mothers to return for vaccination appointments, but due to being overburdened with service provision, we are unable to initiate vaccinations during these focused PNC visits…Consequently, home delivery and missed postnatal follow-up contribute to gaps in vaccine schedule and benefit information for these mothers.”*

In line with previous studies^(23,24)^, the current study revealed that children with mothers/caregivers who had poor knowledge about vaccination schedules increased the odds of being non-vaccinated and incompletely vaccinated by 5.54 and 3.30 times compared with mothers/caregivers having good knowledge, respectively. This may be explained by; mothers who had awareness about the vaccination schedule and might take their children for vaccination based on schedule. The finding implies that interventions in the awareness creation of mothers/caregivers about vaccination schedule will encourage parents to fully vaccinate their children, and monitor whether vaccination has been completed or incomplete^(25–27)^..

*“…We vaccinate our children when health extension workers come to give vaccine through home to home or when we take our children to the health center for treatment purposes because as you know we are not educated, or we have no information about vaccination schedule. If they advise on when and why to vaccinate, I can follow exactly what they advised me”.* This finding supports the finding of local study results, we do have a problem with providing quality health information that can change our clients. The information we deliver does not cast out the problem with the vaccine schedule and does not change behavior. In line with a study conducted in Ethiopia^(28)^.

Similarly, the odds of non-and incomplete vaccination are increased for children with mothers/caregivers who had poor knowledge about vaccine VPDs by 2.78 and 2.71 folds, respectively. The current finding is supported by the result of a study conducted in another place ^(24,25,29,30)^. In addition, some studies show children of mothers who knew that vaccination is used to prevent VPDs were more likely to be fully vaccinated^(20,27)^. This might be due to that mothers/caregivers who were aware of some basic facts about VPDs may be concerned that their child can get the disease if not vaccinated ^(27,31)^. This result indicates that interventions aimed at improving mothers’/caregivers’ knowledge about VPDs increase the likelihood of vaccinating their children.

The present study also revealed that children with mothers/caregivers who had negative attitudes towards vaccination have a significant association with non- and incomplete vaccination status. Similarly, a study conducted in Cameroon revealed that children with mothers/caregivers having negative attitudes towards vaccination increase the likelihood of non-vaccination ^(32)^. This may be explained by the fact that mothers who had a misperception and belief about vaccination and its side effects had a high chance of incomplete and/or non-vaccinating their children. The finding implies that discussion with mothers/caretakers about the benefits of vaccination can change their attitude towards the vaccine.

> *Supported by qualitative findings… "After vaccination, my child developed a high fever and cried throughout the night. This experience worried many mothers in our kebele, including myself*. *…Maal …. As a result, I’m hesitant to bring my child in for the next round of vaccinations. When my child has these symptoms, we don’t know what to do or what home remedies might help."*

In line with previous studies ^(20)^ ^(21)(33)^, the present study also revealed that the absence of a nearby health facility was found to be an important predictor for non-vaccination of children. This could be due to those mothers/caregivers who travel long distances to vaccinate their children might not be encouraged to take their children for vaccination. It implies that strengthening interventions such as outreach EPI programs is essential to identify and address those children who have not started vaccination yet.

> Also finding qualitative supports “…. *We identified the Abayi area for outreach but know we do not provide outreach service in the identified area because the current security situation is not good and travelling with a motor bicycle costs around 400 ETB. It is impossible to provide outreach service like before and we appoint them to vaccinate their child at an adjacent health facility but they didn’t come “*.

This study also revealed that transportation incurred cost has a positive association with non-vaccination of children aged 12-23 months. This study also revealed that transportation incurred cost has a positive association with non-vaccination of children aged 12-23 months. This finding is comparable with previous studies in which vaccination status is significantly associated with household income^(34)^ ^(35)^. Although vaccination services are offered free in Ethiopia, the time and financial cost of reaching the health facilities that provide the vaccination services may be an obstacle for parents to initiate vaccinating their children.

> *Supported by …. “We can’t travel two-three hours for vaccination, here I have 3 children none of them were vaccinated because both health post and health center are very far to get vaccination service”*.

In this study, being from mothers/caregivers who had negative perceived benefits of the vaccine were more likely to be non-vaccinated than fully vaccinated compared to children from mothers/caregivers who had positive perceived benefits of the vaccine. This finding is in line with the result of studies conducted in Rwanda, which show negative perception about the benefits of vaccination increases the chance of childhood vaccination^(36)^ and hinders mothers from vaccinating their children^(32)^. Furthermore, having a positive perception of vaccination was strongly associated with full vaccination of the children in a study conducted in Serbia ^(37)^. This, finding shows that there is a gap in promoting the benefits of childhood vaccination and advising mothers/caregivers about the importance of vaccination during their perinatal period. Thus, we encourage healthcare providers to plan to provide information about the benefits of vaccines to mothers/caregivers.

Lastly, the provision of counselling about vaccination by health workers was significantly associated with incomplete vaccination of children aged 12-23 months. As mentioned in the earlier findings in this study, the mothers/caregivers’ poor knowledge about VPDs and vaccination schedule, and their attitude towards vaccines were indicated as poor and negative in this study, respectively. This might be due to a lack of counselling from healthcare workers and/or inappropriate communication between healthcare providers and mothers/caregivers while providing counselling on childhood vaccination. Therefore, it is suggested that healthcare providers and stakeholders need to consider developing and implementing an effective communication approach while counselling about vaccination.

### Strengths and Limitations of the Study

In this study, the outcome variable was assessed into a three-way category, which addresses the associated factors of non-vaccinated and incompletely vaccinated separately using fully vaccinated as the reference category. Thus, it is very helpful to develop and implement specific interventions tailored to each category. However, the findings of the current study should be interpreted cautiously for the following reasons. First, mothers/caregivers who did not have child immunization cards were asked to recall the vaccine that the children received, which may introduce recall bias. However, it has been reported that in nations where immunization records are not available maternal recall provides a valid estimate of vaccination coverage. Second, this study is not free from social desirability biases on child vaccination status and it may overestimate the findings. Finally, due to the cross-sectional nature of the study design, data on vaccination status and certain factors were collected at the same time. Thus, it may be difficult to develop a causal relationship between vaccination status and identified factors.

## Conclusion

Despite the health and economic benefits of childhood vaccination, the present study found that a significant proportion of children were non-vaccinated and incompletely vaccinated in the study area. Our study also revealed that full vaccination coverage was below the governmental plan of 90% in 2020 and the 90% benchmark target set by the WHO on the vaccination schedule for reducing childhood illness and death.

After controlling for potential confounding factors, the multinomial logistic regression result showed that place of delivery, maternal/caregivers’ knowledge about VPDs and vaccine schedule, and their attitude towards vaccination were significantly associated with non- and incomplete vaccination status. In addition, nearby health facilities, and transportation incur costs and mothers/caregivers’ perceived benefits of vaccine were significantly associated with non-vaccination. Moreover, the study identified that lack of counselling about vaccination is positively associated with incomplete vaccination. As a result, the investigators recommend narrowing the observed variation in child vaccination by designing and implementing interventions that address the identified factors for non-and incomplete vaccination.

Particularly, the use of locally available structures like the Health Development Army (HDA) and health extension workers to provide health education to parents about VPDs and vaccine schedules needs to be strengthened. Likewise, efforts to increase maternal health service utilization such as institutional delivery and provision of counseling on child vaccination are focus areas when designing public health interventions to improve vaccination status. In addition, the issues of transportation cost and inaccessibility of health facilities that provide EPI service can be addressed by strengthening outreach EPI programs and/or increasing the number of new vaccination sites/clusters. Finally, strategies to trace defaulters are very crucial to addressing those children who were not fully vaccinated.

## Data Availability

all required data is available and submitted separately

## List of abbreviations

AOR: Adjusted Odds Ratio
BCG: Baccil Calmette Guerin
CI: Confidence Interval
EPI: Expanded program on Immunization
VPDs: Vaccine Preventable Diseases
WHO: World Health Organization

## Declarations

### Ethics approval and consent to participate

Ethical approval was obtained from Jimma University’s ethical review committee with committee’s reference number **IRB-00018/2020** submitted to the East Bale Zone Health Department. An official support letter was written by the Zonal Health Department and given to the Gindhir Health office. After explaining the study’s purpose, procedure, potential benefits and risks, and their right to participate or not, written informed consent was obtained from mother / care giver of the study participants. Data were collected anonymously; information obtained from study participants was used for study purposes only and not disclosed to ensure confidentiality. Those mothers/caretakers with children vaccination status non-and incompletely vaccinated were advised to vaccinate their children. Furthermore, general information on the importance of child vaccination was provided for mothers/caregivers with children of non-and incomplete vaccination.

### Consent for publication

Not applicable.

### Availability of data and materials

The datasets used and/or analyzed during the current study are available from the corresponding author on reasonable request

### Authors’ contributions

G.T. conceptualized the investigation and methodology, participated in data collection, conducted data entry, analyzed the data, interpreted the result, wrote the original paper, and prepared the manuscript. D.H., A. L. and T. Y contributed to the conceptualization of the study and methods, data analysis, and result interpretation and reviewed the manuscript. A. Z and A.W critically review the manuscript.

## Acknowledgements

First, we would like to say thanks to God. The authors also express their deepest gratitude Ethiopian Field Epidemiology and Laboratory Training Program (EFELTP)and the Department of Epidemiology, Faculty of Health, Jimma University. Our gratitude also goes to the East Bale zone health department and Gindhir rural district health office. Finally, we would like to expand our thankfulness to study participants, supervisors and data collectors.

